# Pain Treatment Strategy and Readmission Rates for Medicare Beneficiaries Post-Acute Ischemic Stroke

**DOI:** 10.1101/2025.09.25.25336638

**Authors:** Madhav Sankaranarayanan, Rebeka Bustamante Rocha, Julianne D. Brooks, Maria A. Donahue, Shuo Sun, Sonia Hernandez Diaz, Alexander Tsai, Joseph P. Newhouse, Sebastien Haneuse, Lidia M.V.R. Moura

## Abstract

**Purpose:** Stroke is highly prevalent and commonly presents with pain. Primary care providers generally manage follow-up care, although ideal pain management strategies remain unclear. Treatment options include gabapentinoids, tricyclic antidepressants, and various antiseizure medications. We aim to analyze differences in hospital readmissions, a quality metric, for those initiating gabapentin in contrast to other medicines for post-stroke pain in older adults.

**Methods:** In this matched cohort study, we analyzed a 20% sample of U.S. Medicare beneficiaries aged 65 and over hospitalized for acute ischemic stroke (AIS) between December 31, 2016, and December 31, 2021, who were discharged home. Individuals met insurance coverage criteria and did not take pain medications before hospitalization. We matched individuals on days from discharge to medication initiation. Individuals who initiated Gabapentin within 90 days of discharge (N = 1,546) were matched to individuals who initiated first-line pharmacological treatments for nerve pain other than Gabapentin within 90 days of discharge (N = 285). We investigated the time to re-admissions using a semi-competing risk framework.

**Results:** The matched cohort of 1,831 initiators had a median age of 76 (IQR 11) and was 57.2% female and 81.3% Non-Hispanic White. Using the semi-competing risk framework, the hazard of readmissions, given that death had not occurred, was not different for Gabapentin initiators, compared to if they had initiated other medications, hazard ratio 0.871 (95% CI: 0.517, 1.466).

**Conclusion:** We found no significant difference in hospital readmission rates between gabapentin and other post-stroke pain treatment strategies. Our findings contribute to the pharmacosurveillance of gabapentin in real-world Medicare beneficiaries.

## INTRODUCTION

Post-stroke pain is a frequent and debilitating complication of stroke, and approximately 1 in 10 patients develops new chronic pain after non-severe acute ischemic stroke (AIS).^1,2^ Although common, it is often underdiagnosed and undertreated, particularly among patients with cognitive or speech impairments affecting symptom reporting.^3^ Post-stroke pain syndromes include those of nociceptive origin (musculoskeletal, shoulder pain, and spasticity-related), neuropathic origin (central post-stroke pain (CPSP)), and headache.^4,5^ Pre-existing chronic pain is also common in this population, particularly in older adults with multiple comorbidities, and many patients present with mixed pain types, further complicating diagnosis and treatment.^4^

Pain management is typically conducted in the outpatient setting by primary care providers. However, there is a paucity of guidelines, and existing recommendations vary in ranking the ideal pharmacological strategy. The American Stroke Association outlines the management for CPSP, recommending amitriptyline and lamotrigine as first-line options.^6^ Second-line medications include pregabalin, gabapentin, carbamazepine, and phenytoin.^7,8^ In contrast, guidelines focused on broader neuropathic pain point to pregabalin and gabapentin as first-line, alongside serotonin and norepinephrine reuptake inhibitors (SNRIs), tricyclic antidepressants (TCAs), and topical analgesics.^9^

Although commonly recommended, gabapentin use in neuropathic pain remains off-label, as FDA approval is limited to adjunct treatment of partial-onset seizures and post-herpetic neuralgia.^10^ Yet, gabapentin use in the U.S. has substantially increased over the past decades, and it was ranked as the 6^th^ most dispensed drug in 2018, with 95% of prescriptions estimated for off-label treatment of chronic pain.^11,12^ Nonetheless, use in older adults remains controversial given their heightened susceptibility to side effects, including delirium, cognitive impairment, increased risk of dementia, dizziness, and falls.^13–15^

These side effects are particularly concerning in vulnerable populations such as older stroke survivors, as they may contribute to poor outcomes, including increased readmissions, morbidity, and mortality. Hospital readmission is a recognized marker of health care quality and an important policy measure.^16^ Considering the high prevalence of stroke, the complexity of post-stroke pain, and the lack of clear treatment guidelines, we aim to compare the safety of pain management strategies after stroke in Medicare beneficiaries aged ≥ 66, using hospital readmissions as a safety outcome.

## METHODS

This study was approved by the Mass General Brigham Institutional Review Board and followed the Strengthening the Reporting of Observational Studies in Epidemiology (STROBE) reporting guidelines.^4^ The requirement for informed consent was waived in our study as we performed a secondary analysis of data routinely collected for billing. The data supporting this study’s findings were collected by The Centers for Medicare & Medicaid Services (CMS) and were made available by CMS with no direct identifiers.^17^ All results were aggregated following CMS Cell Suppression Policies. Restrictions apply to the availability of these data, which were used under license for this study. Medicare data are available through CMS with their permission. We included the code that produced the Supplemental Materials (Statistical Code) findings.

### Study Design

We established the criteria for selecting a retrospective cohort of beneficiaries hospitalized with AIS using a 20% random sample of Traditional Medicare Claims. We identified 92,852 patients who were admitted for AIS between December 31^st^, 2016, and December 31^st^, 2021, and with at least one year of enrollment in the Traditional Fee-For-Service Medicare Parts A (hospital insurance information), B (preventative and medically necessary services or medical insurance information), and D (prescription drug coverage information) before AIS hospitalization. We used acute hospitalization records from the Medicare Provider Analysis and Review (MedPAR) file. AIS hospitalizations were selected based on the principal AIS ICD-10 code I63, a validated method to capture AIS using administrative claims.^18^ We took the first hospitalization chronologically for beneficiaries with more than one AIS hospitalization.

We included patients aged 66 and above (a population that is more vulnerable to adverse outcomes due to frailty and multiple comorbidities),^19^ admitted to an acute hospital, and discharged home (short-stay hospitalization). We excluded patients with a recorded AIS diagnosis 12 months before hospitalization and patients with one or more recorded outpatient prescriptions for any medications recommended for CPSP^6^ within six months before admission.

### Baseline Characteristics

For each beneficiary, we captured demographic characteristics, i.e., age, documented sex, reported race/ethnicity, dual Medicare and Medicaid eligibility status, US region of hospitalization, and the original reason for Medicare entitlement provided in Medicare’s Master Beneficiary Summary (MBSF) File. We used the self-reported race/ethnicity variable provided in MBSF (White, Black, Hispanic, Asian, American Native, other, unknown).^7^

We also obtained baseline stroke severity through modified Rankin Scores (mRS).^20^ The mRS has seven total categories ranging from no or low disability to death: 0 (no symptoms), 1 (no significant disability), 2 (slight disability), 3 (moderate disability), 4 (moderate to severe disability), 5 (severe disability), 6 (death).^21^ This study used a validated claims-based algorithm to classify mRS as a binary outcome, grouping scores 0 to 3 into minor disability and 4 to 6 into moderate to severe disability.^22^ The algorithm could accurately identify disability status with an ROC AUC of 0.85.^22^ As an additional measure of stroke severity at baseline, we used the length of stay (LOS) during AIS admission.

We analyzed comorbid conditions present in the 12 months before stroke admission. We used inpatient and outpatient diagnosis codes in the Inpatient, Outpatient, and Carrier Medicare Claims files. We examined pre-admission history of myocardial infarction (MI), congestive heart failure (CHF), peripheral vascular disease (PVD), cardiovascular disease (CVD), chronic obstructive pulmonary disease (COPD), paralysis, diabetes, renal disease, liver disease, rheumatism, and Alzheimer’s disease and related dementia (ADRD) for each beneficiary using the ICD-10 diagnosis codes listed in the Supplementary Materials (**Table S1**) and the National Cancer Institute’s Surveillance, Epidemiology, and End Results (SEER)-Medicare: Comorbidity SAS Macros.^14,15^ This algorithm required diagnoses (not just specific codes) in outpatient claims to appear in at least two different claims more than 30 days apart. Additionally, the MBSF includes death dates.

### Treatment Strategy

We identified patients who initiated medications for post-stroke pain within 90 days post-discharge. The medication initiators were defined as the beneficiaries with an initiation event of medications recommended for CPSP within 90 days of post-stroke discharge, beginning at the index acute hospitalization discharge date. We used an intention-to-treat strategy, so we categorized individuals based on whether they were prescribed a drug, indicated by a prescription claim for gabapentin, amitriptyline, lamotrigine, pregabalin, carbamazepine, or phenytoin. A complete list of brand and generic medication names included in our analysis is provided in **Supplemental Table S2**. The final eligible sample included 1,831 patients (**Figure 1**). We grouped initiators into two groups: gabapentin initiators and other medication initiators (including amitriptyline, lamotrigine, pregabalin, carbamazepine, and phenytoin). We followed individuals from the medication initiation date. The time zero of our analysis is the time of this initiation event.

**Figure 1.**
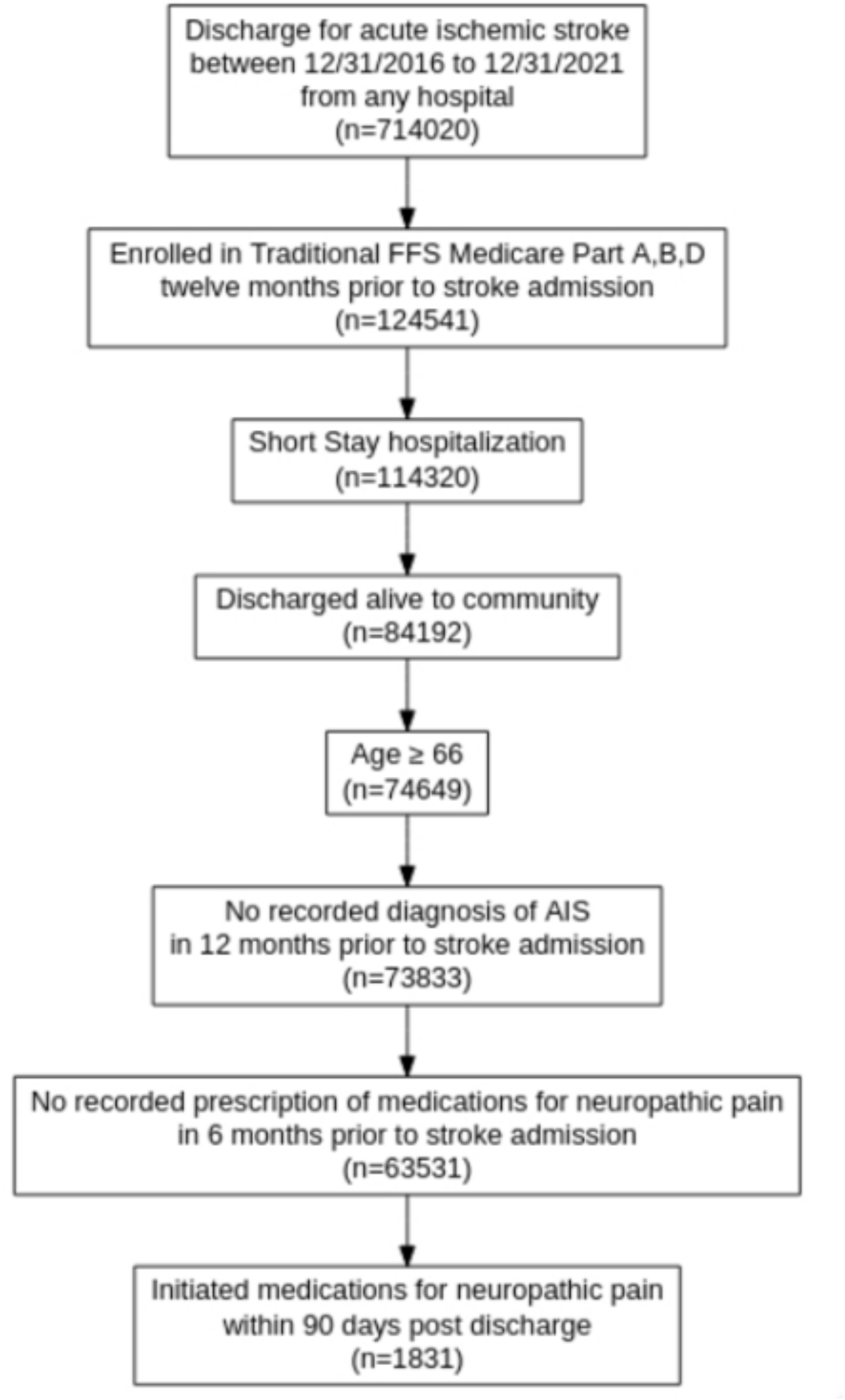
Schematic Describing Eligibility Criteria. **Legend:** Describes the sampling process resulting in a sample of 1831 patients. Medicare data files used: MedPAR (Inpatient data), MBSF (Summary data); Diagnosed for Acute Ischemic Stroke based on ICD-10 codes (I63 and I63.9); Part A: Hospital Insurance, Part B: Medical Insurance; Part D: Drug coverage. AIS, Acute Ischemic Stroke; FFS, Fee-for-Service.

We implemented a matched cohort design, allowing us to address confounding bias by balancing the covariates included in the matching process. Matching is coupled with an analytic, regression-based adjustment for remaining (observed) confounding. We have misaligned treatment starts in our setting, a common issue in real-world time-to-event analysis.^16^ We matched groups on days to treatment initiation. We include figures on the distribution of this time to initiation in the Supplementary Materials (**Figure S1**). This approach can help mitigate residual selection bias due to time-varying covariates that change between discharge and initiation, ensuring a valid comparison between the two groups during a similar risk period.

We used two matching strategies for our cohort: exact matching on time to initiation, and nearest matching using time to initiation and other baseline covariates. For the precise matching, we find weights for every gabapentin initiator, such that the distribution of time to initiation is the same in both groups. For the nearest matching, we identified five matches for each beneficiary using other medications in the gabapentin treatment group. We calculated the Mahalanobis distance from other medication initiators to gabapentin initiators based on days from discharge to medication initiation, sex, and race (White vs non-White). We predicted mRS (which we describe in baseline characteristics), and selected matches based on the shortest distance.^23^ In this matching process, patients on gabapentin were used as controls for each patient utilizing another medication (matching without replacement). We used the exact matching cohort in subsequent analyses and included the results of the nearest matching, including the complete cohort analysis, in the Supplementary Materials (**Table S3**).

### Outcomes

Our primary outcome is the time to hospital readmissions, with a follow-up period of 180 days after initiation. Hospital readmissions are an essential quality metric for healthcare systems and CMS, especially in the older adult population.^24,25^ Mortality is an important consideration when analyzing hospital readmissions, as individuals may die within the observation period before readmission. Mortality acts as a competing risk, and we also measured time to a mortality event for the cohort. We identified hospital readmissions using acute hospitalization claims occurring after the index stroke hospitalization in the MedPAR file. We used the beneficiary’s date of death [BENE_DEATH_DT] from the Medicare MBSF, which comes from several sources, including the Social Security Administration. Overall, 99% of the death information in the MBSF has been validated.^19^ Individuals were followed from the day of medication initiation to the first occurrence of the outcome, mortality, or a censoring event (such as the end of the study observation period). Using Medicare claims data, we followed individuals to the end of the study period.

### Statistical Analysis

Our primary statistical goal was to investigate whether initiating gabapentin (in contrast to initiating other medications for CPSP) post-stroke affects readmission rates. We used a semi-competing risks framework to analyze hospital readmission with mortality as a competing risk. The semi-competing risks framework^16^ allowed us to consider the occurrence of a non-terminal event (readmission), which is subject to a terminal event (death). We used an illness-death model using a Weibull distribution, which defines three hazard functions. The first hazard function is for readmissions, given that neither readmissions nor death has occurred. The second hazard function is for death, given that neither readmissions nor death has occurred. The third function is for the time to death after a readmission event. We presented results for the Weibull illness-death model based on the semi-Markov specification. We used the SemiCompRisk and SemiCompRiskFreq packages for R.^26^

We presented two model formulations for adjustment. The first model includes potential confounding variables selected based on preplanned analysis. These are age, sex, race, ADRD, predicted mRS, and time to initiation. The second model includes all covariates of the first model, and additional variables have a substantial difference between the two initiation groups, given by a standardized mean difference (SMD) above 0.1. These are all the covariates from the first model, along with length of hospitalization, US region of hospitalization, history of cardiovascular disease, and ulcers. We included time to initiation in our covariates despite performing matching on the same variable. Prior work has shown that any inadequacies in reducing selection bias in the matching stage can be further reduced using the same variables in the regression stage.^27^ We focused on using the first model for our analysis, and we included the results for the second model and an unadjusted model in the Supplementary Materials.

### Secondary Pre-planned Stratified Analyses

In addition to the primary analysis, we completed stratified analyses based on age (below 76 years of age, from 76 to 85 years of age, above 85 years of age), ADRD status (with baseline ADRD and without baseline ADRD), and mRS (>4 and ≤4).

## RESULTS

### Study Population Characteristics

We examined a sample of 1,831 beneficiaries discharged for an AIS during the study period (**Figure 1**), where 1,546 (84.43%) initiated gabapentin within 90 days post-discharge, and 285 (15.57%) initiated other medications for CPSP within 90 days post-discharge. Sample characteristics are presented in **Table 1**, and in **Table S3** for the matched cohort (using nearest matching). The median age was 76 (IQR 11), with 57.2% female, 81.3% White, 10.5% Black/African American, 2.4% Hispanic, and 2.4% Asian beneficiaries (Table 1). Using a 0.1 threshold for defining a substantial difference based on standardized mean differences, ADRD, cardiovascular disease (CVD), and ulcers at baseline were associated with the initiation strategy. Among gabapentin initiators, 2.7% had ADRD at baseline, and among other initiators, 2.7% had ADRD at baseline (SMD 0.16). CVD was present at baseline in 9.8% of gabapentin initiators and 14.7% of ASM initiators (SMD 0.15). We present the frequency of other baseline comorbidities in Table 1. We found that the length of hospitalization stay (in days) and the predicted mRS, representing two measures of stroke severity, were associated with initiation strategy. The mean length of stay was 4.1 days (SD 3.8) for gabapentin initiators and 3.7 days (SD: 3.1) for other pain medication initiators (SMD: 0.12). More gabapentin initiators had a predicted mRS indicating severe disability at baseline. The predicted mRS was ≥4 in 71% of gabapentin initiators and 64% of other medication initiators (SMD: 0.16).

**Table 1.** Characteristics of Patients Stratified by Initiation Strategy.

| Characteristics | Overall Sample | Initiation Strategy |  | SMD |
| --- | --- | --- | --- | --- |
|  |  | Other (n=285) | Gabapentin (n=1546) |  |
| Age (median (IQR)) | 76 (11) | 76 (11) | 76 (11) | 0.02 |
| Age Categories (%) |  |  |  | 0.02 |
| 66-75 | 788 (43.0) | 126 (44.2) | 662 (42.8) |  |
| 76-85 | 727 (39.7) | 110 (38.6) | 617 (39.9) |  |
| 86+ | 315 (17.2) | 49 (17.2) | 266 (17.2) |  |
| Sex (%) |  |  |  |  |
| Male | 784 (42.8) | 124 (43.5) | 660 (42.7) | 0.02 |
| Female | 1047 (57.2) | 161 (56.5) | 886 (57.3) |  |
| Race/Ethnicity (%) |  |  |  |  |
| White | 1488 (82.9) | 237 (83.2) | 722 (87.4) | 0.15 |
| Black | 192 (10.5) | 26 (9.1) | 166 (10.7) |  |
| Asian | 44 (2.4) | <11 (1.8) | >33 (2.5) |  |
| Hispanic | 44 (2.4) | <11 (2.5) | >33 (2.4) |  |
| American Native | <11 (*) | <11 (*) | <11 (*) |  |
| Other | 24 (1.3) | <11 (1.4) | >13 (1.3) |  |
| Unknown | 29 (1.6) | <11 (2.1) | >18 (1.5) |  |
| US Regions (%) |  |  |  |  |
| Midwest | 402 (23.7) | 65 (24.7) | 337 (23.5) | 0.29 |
| Northeast | 367 (21.6) | 44 (16.7) | 323 (22.5) |  |
| Southeast | 453 (26.7) | 92 (35.0) | 361 (25.2) |  |
| Southwest | 211 (12.4) | 36 (13.7) | 175 (12.2) |  |
| West | 263 (15.5) | 26 (9.9) | 237 (16.5) |  |
| Year of Discharge (%) |  |  |  |  |
| 2017 | 435 (23.8) | 70 (24.6) | 365 (23.6) | 0.09 |
| 2018 | 375 (20.5) | 50 (17.5) | 325 (21.0) |  |
| 2019 | 338 (18.5) | 55 (19.3) | 283 (18.3) |  |
| 2020 | 362 (19.8) | 58 (20.4) | 304 (19.7) |  |
| 2021 | 320 (17.5) | 52 (18.2) | 268 (17.3) |  |
| Baseline ADRD (%) | 59 (3.2) | 17 (6.0) | 42 (2.7) | 0.16 |
| Acute MI (%) | 39 (2.1) | <11 (2.5) | >28 (2.1) | 0.03 |
| History of MI (%) | 45 (2.5) | <11 (2.5) | >34 (2.5) | <0.01 |
| CHF (%) | 209 (11.4) | 28 (9.8) | 181 (11.7) | 0.06 |
| PVD (%) | 194 (10.6) | 35 (12.3) | 159 (10.3) | 0.06 |
| CVD (%) | 193 (10.5) | 42 (14.7) | 151 (9.8) | 0.15 |
| COPD (%) | 231 (12.6) | 38 (13.3) | 193 (12.5) | 0.03 |
| Hemiplegia/Paraplegia (%) | 18 (1.0) | <11 (*) | <11 (*) | 0.08 |
| Diabetes (%) | 453 (24.8) | 81 (28.4) | 372 (24.1) | 0.10 |
| Renal Disease (%) | 228 (12.5) | 34 (11.9) | 194 (12.6) | 0.02 |
| Liver Disease (%) | <11 (*) | <11 (*) | <11 (*) | 0.05 |
| Ulcers (%) | 18 (1.0) | <11 (2.1) | >7 (0.8) | 0.11 |
| Rheumatic Disease (%) | 67 (3.7) | 11 (3.9) | 56 (3.6) | 0.01 |
| Predicted mRS Categories (%) |  |  |  |  |
| ≤ 4 | 553 (30.2) | 104 (36.5) | 449 (29.1) | 0.16 |
| > 4 | 1277 (69.8) | 181 (63.5) | 1096 (70.9) |  |
| Length of Stay (mean (SD)) | 4.11 (3.73) | 3.74 (3.14) | 4.18 (3.82) | 0.12 |
| 180-day Readmission<sup>†</sup> (%) | 188 (10.3) | 37 (13.0) | 151 (9.8) | 0.10 |
| 180-day Mortality<sup>†</sup> (%) | 190 (10.3) | 36 (12.6) | 154 (10.0) | 0.09 |

**Legend:** Characteristics of patients stratified by initiation strategy within 90 days post-discharge
<sup>†</sup>-180 days post initiation of medication
\*-indicates a percentage that has been obscured for data privacy
IQR, Inter-Quartile Range; ADRD, Alzheimer's Disease and Related Dementia; CHF, Congestive Heart Failure; COPD,
Chronic Obstructive Pulmonary Disease; CVD, Cardiovascular Disease; MI, Myocardial Infarction; PVD, Peripheral
Vascular Disease.

We found no missing demographic data and minimal missingness for baseline medical conditions (4%) for our selected cohort. We assumed no baseline conditions for those without a recorded one. We described the characteristics of eligible patients stratified by initiation strategy in **Table 1**.

### Outcome: Readmission

Among the gabapentin initiators in the exact matching cohort, there were 151 (9.8%) readmission events and 154 (10%) mortality events within 180 days post-initiation. The average follow-up time (to end of the follow-up window or death) was 158.17 days, and the total follow-up time was 757.84 person-years. Also, among the other medication initiators, there were 37 (13%) readmission events and 36 (13%) mortality events within the observation period. We present counts of events in the nearest matching cohort in the Supplemental Materials (**Table S3**). Cumulative incidence curves are shown in the Supplemental Materials (**Figure S2**).

Using exact matching on time to initiation and adjusting for age, sex, race, ADRD, predicted mRS and time to initiation, we found that among patients who initiated gabapentin, the hazard of readmission (given death has not occurred) was 13% lower than if they had initiated other medication for CPSP, hazard ratio 0.87 (95% CI: 0.52, 1.47). This hazard ratio is not statistically significant, but the point estimate is sufficiently far from 1 that we have reported it as such. We included the hazard ratios in **Table 2** and the cause-specific cumulative incidence curves in **Figure 2**.

**Table 2.**
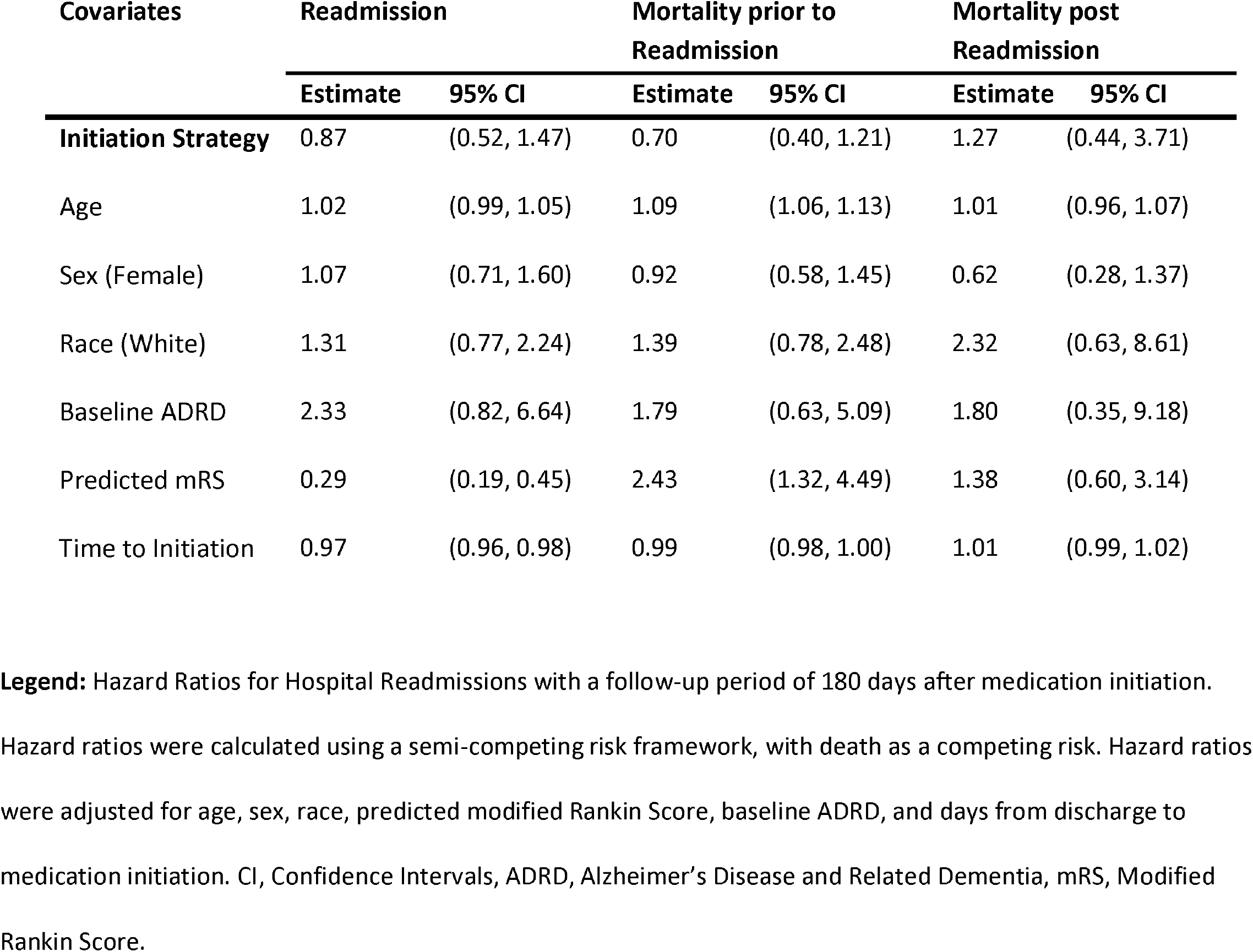
Hazard Ratios of Covariates in the Semi-Competing Risks Model.

| Covariates | Readmission |  | Mortality prior to<br>Readmission |  | Mortality post<br>Readmission |  |
| --- | --- | --- | --- | --- | --- | --- |
|  | Estimate | 95% CI | Estimate | 95% CI | Estimate | 95% CI |
| Initiation Strategy | 0.87 | (0.52, 1.47) | 0.70 | (0.40, 1.21) | 1.27 | (0.44, 3.71) |
| Age | 1.02 | (0.99, 1.05) | 1.09 | (1.06, 1.13) | 1.01 | (0.96, 1.07) |
| Sex (Female) | 1.07 | (0.71, 1.60) | 0.92 | (0.58, 1.45) | 0.62 | (0.28, 1.37) |
| Race (White) | 1.31 | (0.77, 2.24) | 1.39 | (0.78, 2.48) | 2.32 | (0.63, 8.61) |
| Baseline ADRD | 2.33 | (0.82, 6.64) | 1.79 | (0.63, 5.09) | 1.80 | (0.35, 9.18) |
| Predicted mRS | 0.29 | (0.19, 0.45) | 2.43 | (1.32, 4.49) | 1.38 | (0.60, 3.14) |
| Time to Initiation | 0.97 | (0.96, 0.98) | 0.99 | (0.98, 1.00) | 1.01 | (0.99, 1.02) |

**Legend:** Hazard Ratios for Hospital Readmissions with a follow-up period of 180 days after medication initiation.
Hazard ratios were calculated using a semi-competing risk framework, with death as a competing risk. Hazard ratios
were adjusted for age, sex, race, predicted modified Rankin Score, baseline ADRD, and days from discharge to
medication initiation. CI, Confidence Intervals, ADRD, Alzheimer’s Disease and Related Dementia, mRS, Modified
Rankin Score.

**Figure 2.**
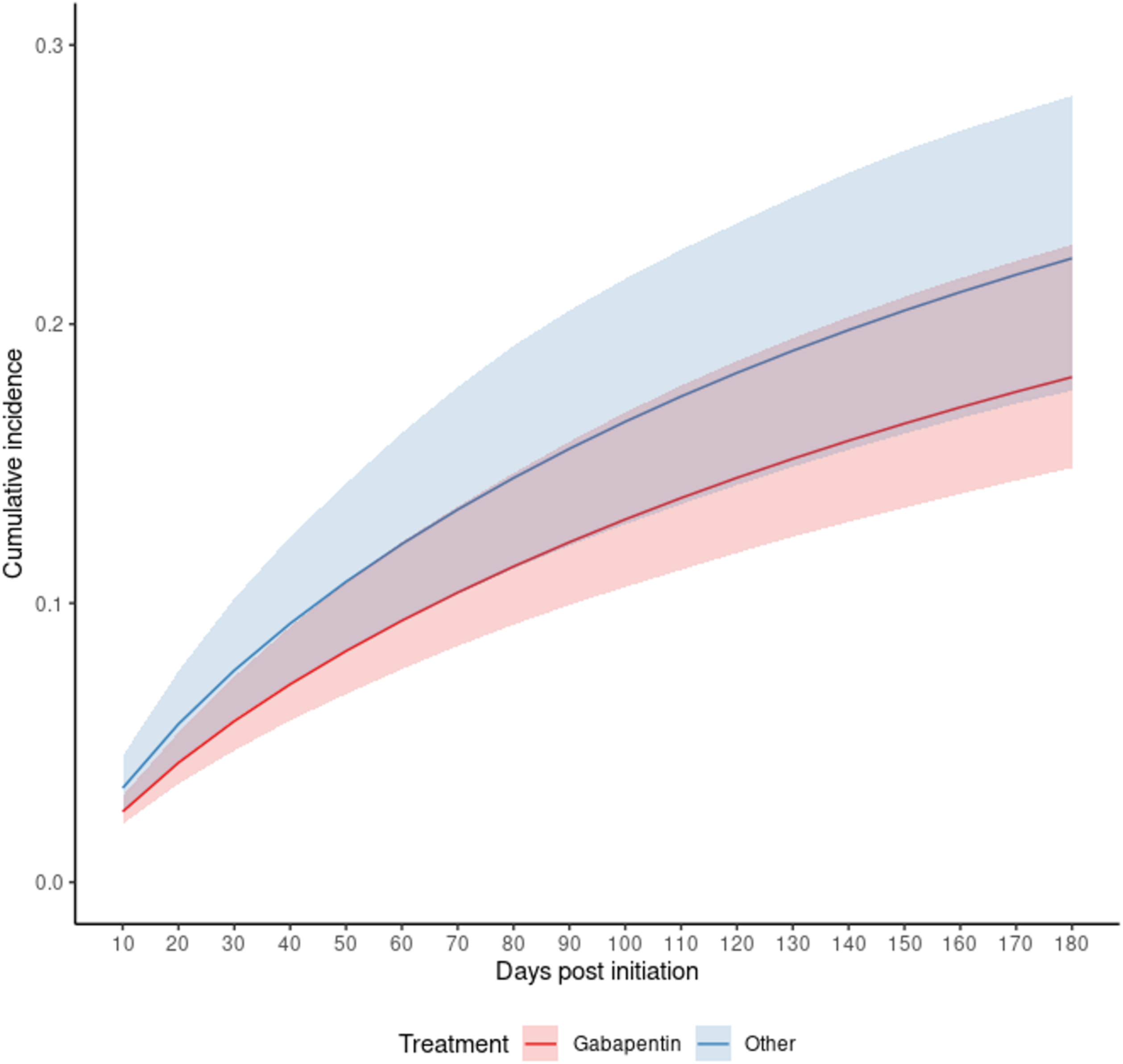
Cause-Specific Cumulative Incidence for Readmission by Medication Initiation Strategy. **Legend:** Cumulative incidence curves based on the semi-competing risks model for all patients. Red: Strategy for gabapentin initiation within 90 days post-discharge from AIS admission. Blue: Strategy for other medication initiation within 90 days post-discharge from AIS admission. Shaded areas: 95% of CIs were constructed using Bootstrap with 500 replications. The confidence bands between the two groups overlap, indicating the difference in incidence between the two groups is not significant.

### Additional Analyses

We included the results of the same analysis conducted on the nearest matching dataset and the whole cohort in the Supplementary Materials (**Table S3**). We include the results of the analysis using the larger model with all variables with an SMD above 0.1, along with an unadjusted model, in the Supplementary Materials (**Table S4**). Additionally, the results for the stratified analyses in the exact matching cohort are presented with the semi-competing risks framework using same model specifications as our primary analysis. It should be noted that due to the small initial sample size, we encountered positivity violations in many of the stratifications. This causes numerical instabilities in the statistical implementations, which means some models do not converge. We have catalogued these analyses in the Supplementary Materials (**Table S5**).

## DISCUSSION

Pain is a common and often debilitating condition after a stroke.^1,4^ Primary care physicians usually manage post-stroke care, since access to specialty neurology care is limited. CPSP guidelines for adults indicate that there is little evidence on the efficacy of treatments,^6,28^ with even less information on older adults. Pain treatment must be carefully managed among older adults due to potential side effects and interactions with other medications.^29,30^ In a cohort of older adults, we analyzed post-stroke pain treatment options and hospital readmissions, a key quality measure. Using a matched cohort design, we found no statistically significant difference in readmissions among Medicare beneficiaries hospitalized for AIS who initiated gabapentin compared to other medications for post-stroke pain. We used a semi-competing risks strategy to account for mortality, strengthening our results.

Gabapentin use has increased substantially over the years across all age groups, despite no studies analyzing long-term safety and effectiveness.^31–33^ Prescriptions tripled from 2.6% in 2002 to 8.4% in 2015,^34,35^ increasing to 9% in 2019-2021.^36^ Several safety concerns have been linked to gabapentin initiation, including increased risk of dementia, falls, and atrial fibrillation among older adults with no pre-existing cardiovascular disease.^14,15,37^

To evaluate safety, we chose readmission as it is an essential marker of healthcare quality and is well-measured in administrative claims data.^16^ Drug-related readmissions are a leading cause of rehospitalization, and up to 9% of older adults experience a drug-related admission, with 22% being preventable.^38^ Beyond the negative impact on patient outcomes and increased morbidity and mortality, readmissions among Medicare beneficiaries are costly and burdensome on the health care system.^39,40^

To use readmission as our key outcome, we must account for patients who die before experiencing a readmission event. We utilize a semi-competing risks framework, which accounts for readmission as a non-terminal event, in contrast to mortality, which acts as a terminal event. Notably, in our study, the point estimate for the hazard ratio indicated a protective effect of gabapentin on readmissions, although it was not statistically significant. Interestingly, recent animal studies^41^ and a small-scale prospective observational study,^28^ discuss potential beneficial effects of gabapentin on post-stroke recovery and CPSP management. Daily administration of gabapentin for six weeks after stroke was shown to improve motor function, and functional recovery persisted even after interruption of the drug.^41,44^ However, other animal studies demonstrate gabapentin insensitivity after long-term use and gabapentin’s selective action on the somatosensory system, hinting at long-term ineffectiveness for pain management.^42,43^ Human subjects’ studies with larger sample sizes are needed to confirm a potential benefit and treatment efficacy.

Additionally, our results show that measures of stroke severity, such as length of stay and predicted mRS, were associated with the medication initiation strategy. Previous studies have pointed to a higher prevalence of post-stroke pain in those with greater stroke severity and prior history of chronic pain.^2,4^ Pain management is, therefore, complex as many patients present with mixed pain types (nociceptive and neuropathic origin). In the population 66 years and older, additional challenges exist, as cognitive and speech impairments affect pain reporting and assessment. Additionally, patients present with multiple comorbidities, decreased renal function, and polypharmacy.^45,46^

### Limitations

The findings for our population sample might not be generalizable to other groups not included in this study, such as Medicare beneficiaries enrolled in Part C (Medicare Advantage, bundled payment insurance coverage options), patients not enrolled in Medicare prescription plans (Part D), or beneficiaries discharged to inpatient rehabilitation units or SNFs. Individuals who are discharged to inpatient or skilled nursing care usually present with variable disabilities and would benefit from assessment in future studies.^47,48^ Lastly, our sample of people with ADRD was too small to complete stratification, as initially planned. The small sample size is likely the cause of the wide confidence intervals reported in our results.

Medicare administrative claims data is collected for billing purposes; therefore, some clinical elements are absent. We could not adjust for factors associated with stroke severity, such as stroke territory. We do not have information on pain severity or pain scores. In addition, prescription claims do not contain information on medication indications, making it challenging to identify if the reason for the prescription was related to the AIS event or concomitant comorbidities. Medications used to treat post-stroke pain are also used to treat other conditions, such as depression, anxiety, mood disorders, and seizures. However, our analysis remains valuable, as Medicare data provides real-world results and it lays the groundwork for future studies that can inform clinical practice and improve health care.^49^

Choosing a matched cohort design for the study methodology reduced the risk of immortal time bias during treatment initiation. Future studies without a matching methodology may find limitations regarding drug initiation time window variation. Additionally, there is always a possibility of unmeasured confounding in our analysis, as we utilized comorbidity information from stroke discharge, but not medication initiation. We used time to initiation, alongside baseline comorbidities, as a proxy variable for any potentially developing comorbidities.

## CONCLUSION

Real-world evidence studies can help provide more information on which pain treatment choice leads to better safety outcomes in older stroke survivors. In our national sample of Medicare beneficiaries hospitalized for acute ischemic stroke and discharged home, there was no difference in readmissions for the gabapentin versus other pain medication strategies. Our findings provide significant contributions to the pharmacosurveillance of gabapentin in older adults following AIS.

## Data Availability

We had a Data Use Agreement approved by the Center for Medicare & Medicaid Services (DUA RSCH-2022-58182). Interested researchers may replicate the study by obtaining the data from CMS. Reproducing this study requires 20% MedPAR, Outpatient, Carrier, and Part D standard analytical files.

## SUPPLEMENTAL MATERIALS

Table S1. ICD-10 diagnosis codes for Baseline Comorbidities

Table S2. Medications Names

Table S3. Nearest Matching Sample Characteristics

Table S4. Comparison of Hazard Ratios for Initiation on Different Cohorts with Different Models

Table S5. Hazard Ratios for Initiation in Stratified Analyses

Figure S1 Histogram of Time to Initiation between Initiation Strategies

Figure S2 Cumulative Incidence Curves with Aalen-Johansen Estimates

Statistical Code

