## Supplementary Materials for "Pain Treatment Strategy and Readmission Rates for Medicare Beneficiaries Post-Acute Ischemic Stroke"

**Included Materials:**

Table S1. ICD-10 diagnosis codes for Baseline Comorbidities

Table S2. Medications Names

Table S3. Nearest Matching Sample Characteristics

Table S4. Comparison of Hazard Ratios for Initiation on Different Cohorts with Different Models

Table S5. Hazard Ratios for Initiation in Stratified Analyses

Figure S1 Histogram of Time to Initiation between Initiation Strategies

Figure S2 Cumulative Incidence Curves with Aalen-Johansen Estimates

Statistical Code

### TABLES

**Table S1. ICD-10 Codes to identify baseline characteristics.**

| Condition | ICD-10 |
|---|---|
| Baseline Dementia | F051, G30, G311 |
| History of MI | I252 |
| CHF | I099, I110, I130, I132, I255, I420, I43, I50, P290, I425-I429 |
| PVD | I70, I71, I731, I738, I739, I771, I790, I792, K551, K558, K559, Z958, Z959 |
| CVD | G45, G46, H340, I6 |
| COPD | I278, I279, J684, J701, J703, J40-J47, J60-J67 |
| Paralysis | G041, G114, G801, G802, G81, G82, G839, G830-G834 |
| Diabetes | E100, E101, E106, E108, E109, E110, E111, E116, E118, E119, E130, E131, E136, E138, E139 |
| Renal Disease | I120, I131, N18, N19, N250, Z940, Z992, N032-N037, N052-N057, Z490-Z492 |
| Liver Disease | I850, I859, I864, I982, K704, K711, K721, K729, K765, K766, K767 |
| Rheumatic Disease | M05, M06, M315, M32, M33, M34, M351, M353, M360 |

**Legend**: Baseline medical conditions were filtered using the following ICD Codes. MI, Myocardial Infraction; CHF, Congestive Heart Failure; PVD, Peripheral Vascular Disease; CVD, Cardiovascular Disease; COPD, Chronic Obstructive Pulmonary Disease

**Table S2. Medication Names**

| | |
|---|---|
| Brand Names | Neurontin, Gabapentin, Gralise, Horizant, Amitriptyline HCL, Perphenazine-Amitriptyline, Chlordiazepoxide-Amitriptyline, Lamotrigine ODT, Lamictal, Lamictal (Orange), Lamictal (Blue), Lamictal XR, Lamictal XR (Orange), Lamictal ODT, Lamictal ODT (Orange), Lamictal (Green), Lamotrigine ER, Lamotrigine, Lamotrigine ODT (Orange), Lamotrigine (Orange), Subvenite (Orange), Subvenite, Subvenite (Blue), Lamotrigine (Green), Lyrica, Lyrica CR, Pregabalin, Tegretol, Tegretol XR, Epitol, Carbamazepine, Carbamazepine ER, Equetro, Carbamazepine XR, Carbatrol, Dilantin, Dilantin-125, Phenytoin Sodium Extended, Phenytek, Phenytoin, Phenytoin Sodium |
| Generic Names | Gabapentin, Amitriptyline HCL, Perphenazine/Amitriptyline HCL, Amitriptyline/Chlordiazepoxide, Amitriptyline HCL/Chlordiazepoxide, Lamotrigine, Pregabalin, Carbamazepine, Phenytoin |

**Legend**: Prescription drug claims were filtered using generic and brand names as displayed in the table.

**Table S3. Characteristics of Patients in Nearest Matching Cohort Stratified by Initiation Strategy**

| | | Initiation Strategy | | |
|---|---|---|---|---|
| **Characteristics** | **Overall Sample** | **Other (n=285)** | **Gabapentin (n=1425)** | **SMD** |
| 180-day Readmission* (%) | 179 (10.5) | 37 (13.0) | 142 (10.0) | 0.095 |
| 180-day Mortality* (%) | 178 (10.4) | 36 (12.6) | 142 (10.0) | 0.084 |
| Age (median (IQR)) | 76 (10.75) | 76 (11) | 75 (11) | 0.016 |
| Age Categories (%) | | | | |
| 66-75 | 712 (41.6) | 126 (44.2) | 586 (41.1) | 0.016 |
| 76-85 | 694 (40.6) | 110 (38.6) | 584 (41.0) | |
| 86+ | 304 (17.8) | 49 (17.2) | 255 (17.9) | |
| Sex (%) | | | | |
| Male | 730 (42.7) | 124 (43.5) | 606 (42.5) | 0.020 |
| Female | 980 (57.3) | 161 (56.5) | 819 (57.5) | |
| Race/Ethnicity (%) | | | | |
| White | 1422 (83.2) | 237 (83.2) | 1185 (83.2) | 0.123 |
| Black | 161 (9.4) | 26 (9.1) | 135 (9.5) | |
| Asian | 31 (1.8) | <11 (1.8) | >20 (1.8) | |
| Hispanic | 38 (2.2) | <11 (2.5) | >27 (2.2) | |
| American Native | <11 (*) | <11 (*) | <11 (*) | |
| Other | 20 (1.2) | <11 (1.4) | >9 (1.1) | |
| Unknown | 29 (1.7) | <11 (2.1) | >18 (1.6) | |
| US Regions (%) | | | | |
| Midwest | 395 (25.0) | 65 (24.7) | 330 (25.0) | 0.269 |
| Northeast | 312 (19.7) | 44 (16.7) | 268 (20.3) | |
| Southeast | 430 (27.2) | 92 (35.0) | 338 (25.6) | |
| Southwest | 200 (12.6) | 36 (13.7) | 164 (12.4) | |
| West | 245 (15.5) | 26 (9.9) | 219 (16.6) | |
| Year of Discharge (%) | | | | |
| 2017 | 402 (23.5) | 70 (24.6) | 332 (23.3) | 0.081 |
| 2018 | 345 (20.2) | 50 (17.5) | 295 (20.7) | |
| 2019 | 317 (18.5) | 55 (19.3) | 262 (18.4) | |
| 2020 | 343 (20.1) | 58 (20.4) | 285 (20.0) | |
| 2021 | 303 (17.7) | 52 (18.2) | 251 (17.6) | |
| Baseline ADRD (%) | 52 (3.0) | 17 (6.0) | 35 (2.5) | 0.175 |
| Acute MI (%) | 36 (2.1) | <11 (2.5) | >25 (2.0) | 0.028 |
| History of MI (%) | 42 (2.5) | <11 (2.5) | >31 (2.5) | <0.001 |
| CHF (%) | 194 (11.3) | 28 (9.8) | 166 (11.6) | 0.059 |
| PVD (%) | 184 (10.8) | 35 (12.3) | 149 (10.5) | 0.058 |
| CVD (%) | 181 (10.6) | 42 (14.7) | 139 (9.8) | 0.152 |
| COPD (%) | 219 (12.8) | 38 (13.3) | 181 (12.7) | 0.019 |
| Hemiplegia/Paraplegia (%) | <22 (0.9) | <11 (*) | 11 (0.8) | 0.088 |
| Diabetes (%) | 417 (24.4) | 81 (28.4) | 336 (23.6) | 0.111 |
| Renal Disease (%) | 213 (12.5) | 34 (11.9) | 179 (12.6) | 0.019 |
| Liver Disease (%) | <11 (*) | 0 (0.0) | <11 (*) | 0.053 |
| Ulcers (%) | 17 (1.0) | <11 (2.1) | >6 (0.8) | 0.112 |
| Rheumatic Disease (%) | 67 (3.9) | 11 (3.9) | 56 (3.9) | 0.004 |
| Predicted mRS Categories (%) | | | | |
| $\leq 4$ | 553 (32.3) | 104 (36.5) | 449 (31.5) | 0.105 |
| $> 4$ | 1157 (67.7) | 181 (63.5) | 976 (68.5) | |
| Length of Stay (mean (SD)) | 4.10 (3.76) | 3.74 (3.14) | 4.17 (3.87) | 0.122 |

**Legend**: Characteristics of patients stratified by initiation strategy within 90 days post-discharge

*-180 days post initiation of medication
IQR, Inter-Quartile Range; ADRD, Alzheimer's Disease and Related Dementia; CHF, Congestive Heart Failure; COPD, Chronic Obstructive Pulmonary Disease; CVD, Cardiovascular Disease; MI, Myocardial Infraction; PVD, Peripheral Vascular Disease.

**Table S4. Comparison of Hazard Ratios for Initiation on Different Cohorts with Different Models**

| **Whole Cohort** | **Readmission** | **Hazard Ratio** | **Lower 95% CI** | **Upper 95% CI** |
|---|---|---|---|---|
| | Unadjusted | 0.676 | 0.412 | 1.109 |
| | Adjusted (Model 1) | 0.809 | 0.517 | 1.266 |
| | Adjusted (Model 2) | 0.838 | 0.500 | 1.405 |
| | **Mortality** | **Hazard Ratio** | **Lower 95% CI** | **Upper 95% CI** |
| | Unadjusted | 0.689 | 0.407 | 1.165 |
| | Adjusted (Model 1) | 0.726 | 0.449 | 1.176 |
| | Adjusted (Model 2) | 0.747 | 0.436 | 1.278 |
| | **Sojourn Time** | **Hazard Ratio** | **Lower 95% CI** | **Upper 95% CI** |
| | Unadjusted | 1.067 | 0.372 | 3.057 |
| | Adjusted (Model 1) | 1.110 | 0.379 | 3.247 |
| | Adjusted (Model 2) | 1.064 | 0.357 | 3.177 |
| **Nearest Matching Cohort** | **Readmission** | **Hazard Ratio** | **Lower 95% CI** | **Upper 95% CI** |
| | Unadjusted | 0.688 | 0.420 | 1.125 |
| | Adjusted (Model 1) | 0.795 | 0.503 | 1.259 |
| | Adjusted (Model 2) | 0.816 | 0.489 | 1.364 |
| | **Mortality** | **Hazard Ratio** | **Lower 95% CI** | **Upper 95% CI** |
| | Unadjusted | 0.682 | 0.404 | 1.152 |
| | Adjusted (Model 1) | 0.699 | 0.426 | 1.147 |
| | Adjusted (Model 2) | 0.708 | 0.413 | 1.212 |
| | **Sojourn Time** | **Hazard Ratio** | **Lower 95% CI** | **Upper 95% CI** |
| | Unadjusted | 1.135 | 0.395 | 3.257 |
| | Adjusted (Model 1) | 1.179 | 0.412 | 3.369 |
| | Adjusted (Model 2) | 1.100 | 0.369 | 3.281 |
| **Exact Matching Cohort** | **Readmission** | **Hazard Ratio** | **Lower 95% CI** | **Upper 95% CI** |
| | Unadjusted | 0.788 | 0.493 | 1.260 |
| | Adjusted (Model 1) | 0.882 | 0.574 | 1.356 |
| | Adjusted (Model 2) | 0.889 | 0.530 | 1.493 |
| | **Mortality** | **Hazard Ratio** | **Lower 95% CI** | **Upper 95% CI** |
| | Unadjusted | 0.665 | 0.401 | 1.102 |
| | Adjusted (Model 1) | 0.702 | 0.436 | 1.130 |
| | Adjusted (Model 2) | 0.716 | 0.414 | 1.238 |
| | **Sojourn Time** | **Hazard Ratio** | **Lower 95% CI** | **Upper 95% CI** |
| | Unadjusted | 1.151 | 0.408 | 3.248 |
| | Adjusted (Model 1) | 1.309 | 0.462 | 3.711 |
| | Adjusted (Model 2) | 1.230 | 0.414 | 3.653 |

**Legend**: Hazard ratios for initiating gabapentin against other medication with a follow-up period of 180 days. We compare the results of the semi-competing risks model for the whole cohort, the exact matching cohort and the nearest matching cohort. Additionally, we compare the unadjusted model with the two adjusted models described in the manuscript.

**Table S5. Hazard Ratios of Initiation Strategy in Stratified Analyses**

| Covariate/Stratum | Readmission | | Mortality prior to Readmission | | Mortality post Readmission | |
|---|---|---|---|---|---|---|
| | **Estimate** | **95% CI** | **Estimate** | **95% CI** | **Estimate** | **95% CI** |
| *Overall* | 0.871 | (0.517, 1.466) | 0.698 | (0.402, 1.213) | 1.274 | (0.438, 3.708) |
| **Age** | | | | | | |
| 66 to 75 years | 0.782 | (0.360, 1.699) | 0.988 | (0.364, 2.681) | 1.706 | (0.293, 9.921) |
| 76 to 85 years | 0.606 | (0.168, 2.191) | 1.276 | (0.365, 4.467) | 0.666 | (0.088, 5.061) |
| 86 years and above | - | - | - | - | - | - |
| **Baseline Dementia** | | | | | | |
| No Dementia | 0.879 | (0.503, 1.535) | 0.703 | (0.390, 1.267) | 1.390 | (0.432, 4.474) |
| Dementia | - | - | - | - | - | - |
| **Predicted mRS** | | | | | | |
| ≤4 (mild-to-moderate) | 0.823 | (0.403, 1.683) | 0.674 | (0.188, 2.421) | 0.363 | (0.087, 1.517) |
| >4 (high) | 0.900 | (0.480, 1.686) | 0.640 | (0.377, 1.085) | 5.623 | (0.533, 5.931) |

**Legend:** Hazard ratios for initiating gabapentin against other medication with a follow-up period of 180 days in different strata of the data. Blank rows imply the semi-competing risks model did not converge.

### FIGURES

**Figure S1. Histogram of Time to Initiation for each Initiation Strategy**

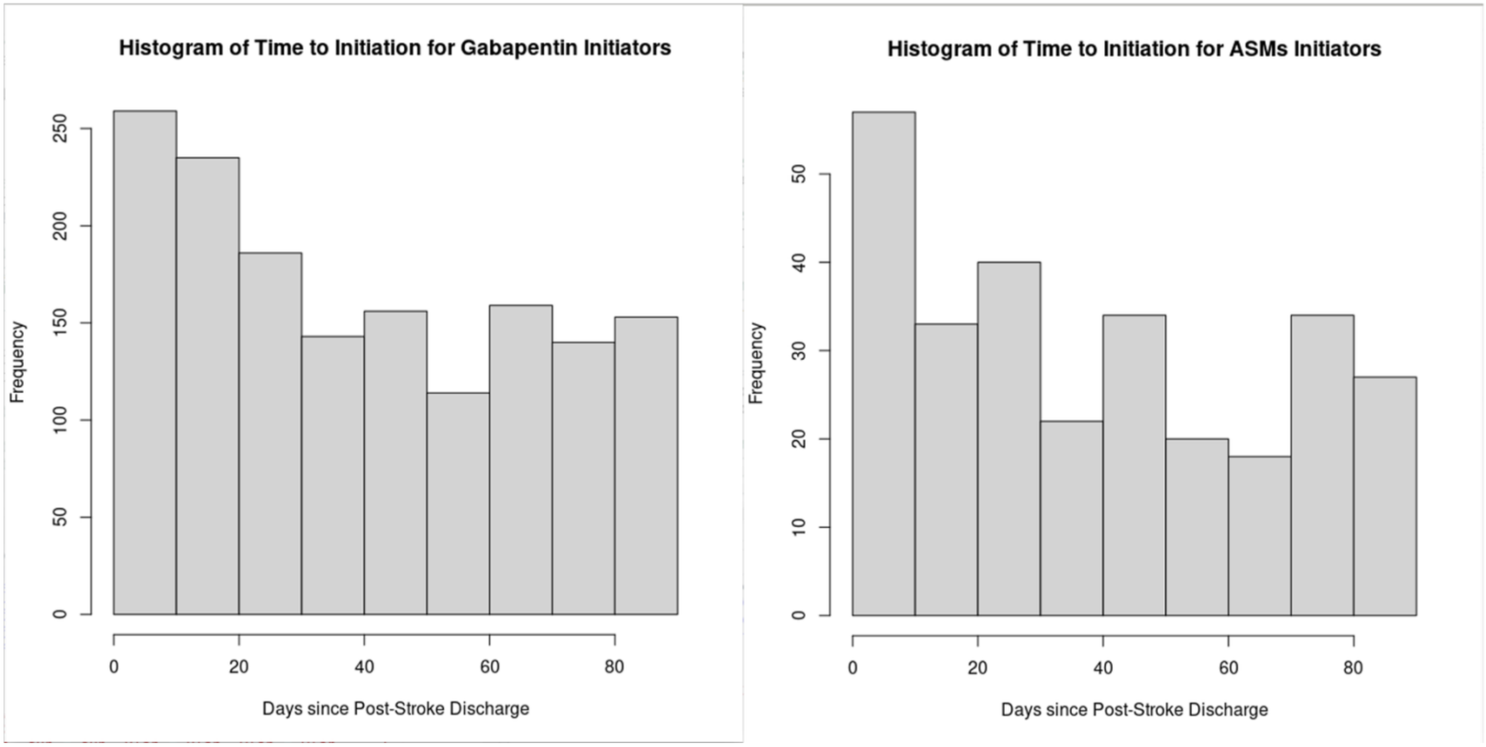


**Legend:** Distribution of days from discharge to medication initiation for patients who initiate gabapentin and patients initiate other medications for pain. We have labelled these as ASMs, but this also includes amitriptyline.

**Figure S2. Aalen Johansen Curves for Readmission and Mortality among the Initiation Strategies**

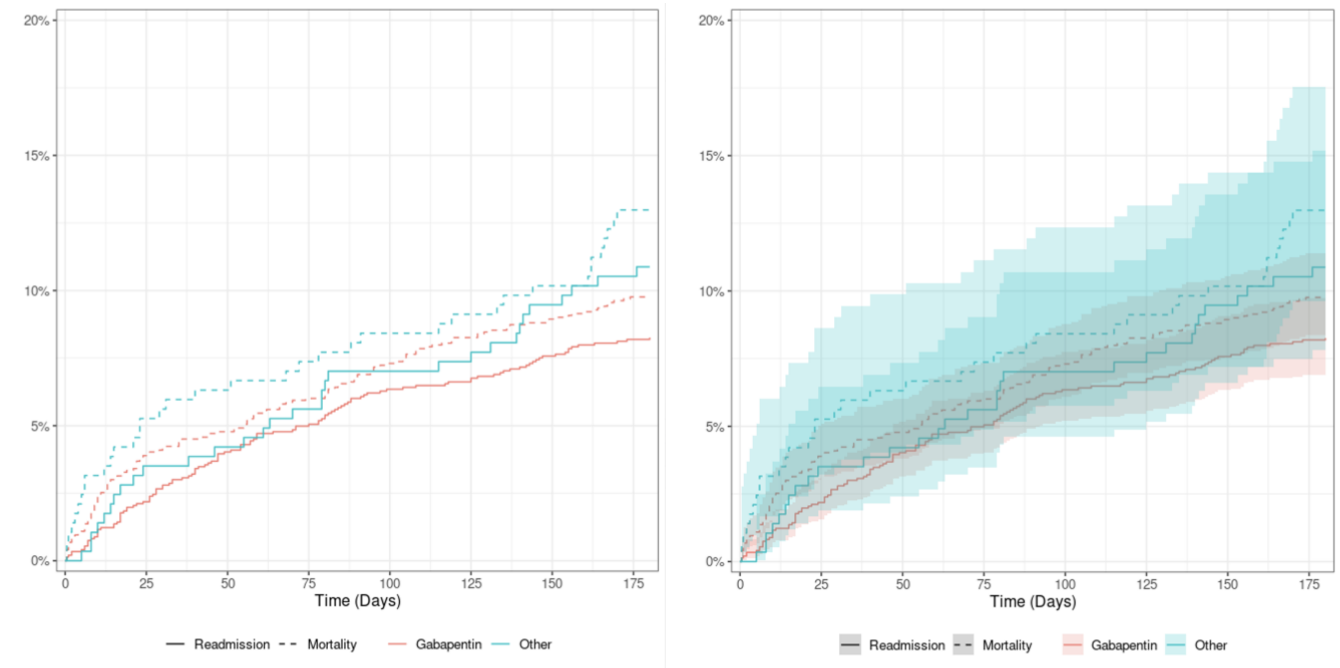


**Legend:** Cumulative incidence curves from the Aalen-Johansen estimates. (Left) Solid lines represent the proportion of individuals with a readmission event at the given time period from the gabapentin initiation group (given in **red**) and the other initiation group (given in **blue**). Dashed lines represent the proportion of individuals with a mortality event at the given time period from the gabapentin and the other initiation groups. (Right) Same figure as the left, with confidence intervals for the Aalen-Johansen estimates.

### STATISTICAL CODE

```
#------Matching Code-----#
library("survival") #package for standard survival analysis
library("SemiCompRisks") #package for semi competing risks analysis
library(dplyr)
library(tidyverse)
library(tidyr)

# install.packages("devtools")
#devtools::install_github("harrisonreeder/SemiCompRisksFreq")
library("SemiCompRisksFreq") #package for semi competing risks analysis - from github

# example code was adapted from SemiCompRiskExamples found on github from:
# https://github.com/harrisonreeder/SemiCompRisksExamples

#### Matching ####
library(MatchIt)


#df_gab is the file with the full cohort, gabapentin is marked with 1, other with 0

df_gab_asm = df_gab_asm %>% mutate(readm_time = case_when(Readm_180_Init==1 ~ readm_day_diff-
dschg_day_diff,
                                            Mort_180_Init==1 ~ death_day_diff-dschg_day_diff,
                                            TRUE ~ 180),
                              mort_time = case_when(Mort_180_Init==1 ~ death_day_diff-dschg_day_diff,
                                           TRUE ~ 180))

##Make race and comorbidities into indicator variables

df_gab_asm = df_gab_asm %>%
  mutate(race_white = as.numeric(race=="White"),
       race_asian = as.numeric(race=="Asian"),
       race_black = as.numeric(race=="Black"),
       race_hispanic = as.numeric(race=="Hispanic"),
       race_native = as.numeric(race=="North American Native"),
       race_other = as.numeric(race=="Other"),
       region_ne = case_when(is.na(USRegions)~0,
                    TRUE~as.numeric(USRegions=="Northeast")),
       dem_ind = as.numeric(dementia=="Yes"),
       cvd_ind = as.numeric(cvd=="Yes"),
       ulc_ind = as.numeric(ulcers=="Yes"))


##Standardize age

df_gab_asm = df_gab_asm %>%
  mutate(age_st = age)
```

```
df_gab_asm$age_st = (df_gab_asm$age_st-80)/10

#Nearest Matching
m.out_asm <- matchit(med_type ~ age + sex + race_white + mrs_predictions + dschg_day_diff, data = df_gab_asm,
            method = "nearest", distance = "mahalanobis", ratio = 5)
#Exact Matching
m.out_asm <- matchit(med_type ~ dschg_day_diff, data = df_gab_asm,
              method = "exact")

##Data frames
df_surv_asm <- match.data(m.out_asm)

##Correcting exceptions (same time of readmission and death)

# #Patients with readmission and mortality on the same day

same_vec = which(df_surv_asm$Readm_180_Init[df_surv_asm$readm_time==df_surv_asm$mort_time]==1)

pat_vec = which(df_surv_asm$readm_time==df_surv_asm$mort_time)[same_vec]

df_surv_asm$mort_time[pat_vec] = df_surv_asm$readm_time[pat_vec]+0.5

##Add indicators for sex


df_surv_asm = df_surv_asm %>%
  mutate(sex_ind = as.numeric(sex=="Female"))

##Adjust invalid survival times

df_surv_asm$readm_time[df_surv_asm$readm_time==0]=0.5

df_surv_asm$mort_time[df_surv_asm$mort_time==0]=0.5


##Cox PH model for mortality


#Unadjusted
model_asm = coxph(Surv(mort_time, Mort_180_Init)~med_type, data = df_surv_asm, weights = weights)

#Model 1
model_asm = coxph(Surv(mort_time, Mort_180_Init)~age + sex_ind + dem_ind + race_white + med_type +
mrs_predictions, data = df_surv_asm, weights = weights)

#Model 2
model_asm = coxph(Surv(mort_time, Mort_180_Init)~age + sex_ind + dem_ind + race_white + med_type +
mrs_predictions + dschg_day_diff + region_ne + cvd_ind, data = df_surv_asm, weights = weights)

summary(model_asm)
```

```
##Cox PH model for readmission

#Unadjusted
model_asm = coxph(Surv(readm_time, Readm_180_Init)~med_type, data = df_surv_asm, weights = weights)

#Model 1
model_asm = coxph(Surv(readm_time, Readm_180_Init)~age + sex_ind + dem_ind + race_white + med_type +
mrs_predictions, data = df_surv_asm, weights = weights)

#Model 2
model_asm = coxph(Surv(readm_time, Readm_180_Init)~age + sex_ind + dem_ind + race_white + med_type +
mrs_predictions + dschg_day_diff + region_ne + cvd_ind, data = df_surv_asm, weights = weights)

summary(model_asm)


##Testing with a semi-competing risks model
#Unadjusted
#form <- Formula::Formula(readm_time + Readm_180_Init | mort_time + Mort_180_Init ~ med_type | med_type
| med_type)
#Adjusted (Model 1)
form <- Formula::Formula(readm_time + Readm_180_Init | mort_time + Mort_180_Init ~ age + sex_ind +
dem_ind + race_white + med_type + mrs_predictions + dschg_day_diff | age + sex_ind + dem_ind + race_white +
med_type + mrs_predictions + dschg_day_diff | age + sex_ind + dem_ind + race_white + med_type +
mrs_predictions + dschg_day_diff)
#Adjusted (Model 2)
#form <- Formula::Formula(readm_time + Readm_180_Init | mort_time + Mort_180_Init ~ age + sex_ind +
dem_ind + race_white + med_type + mrs_predictions + dschg_day_diff + region_ne + cvd_ind | age + sex_ind +
dem_ind + race_white + med_type + mrs_predictions + dschg_day_diff + region_ne + cvd_ind | age + sex_ind +
dem_ind + race_white + med_type + mrs_predictions + dschg_day_diff + region_ne + cvd_ind)


fit_asm = FreqID_HReg2(Formula = form, data = df_surv_asm, model = "semi-Markov", extra_starts = 0, hazard =
"weibull", frailty = TRUE, optim_method = c("BFGS"), weights = df_surv_asm$weights)

summary(fit_asm)

#### Subsetting with age ####

##Comparison between gabapentin and other pain meds (other pain meds are cases)
df_gab_comp_age_1 = df_gab%>%
  filter(med_type>0, age<=75)%>%
  mutate(med_type=1-as.integer(med_type/2))

df_gab_comp_age_2 = df_gab%>%
  filter(med_type>0, age>75, age<=85)%>%
  mutate(med_type=1-as.integer(med_type/2))

df_gab_comp_age_3 = df_gab%>%
```

```
  filter(med_type>0, age>85)%>%
  mutate(med_type=1-as.integer(med_type/2))

df_gab_comp_age_1 = df_gab_comp_age_1 %>% mutate(readm_time = case_when(Readm_180_Init==1 ~
readm_day_diff-dschg_day_diff,
                                              Mort_180_Init==1 ~ death_day_diff-dschg_day_diff,
                                              TRUE ~ 180),
                                 mort_time = case_when(Mort_180_Init==1 ~ death_day_diff-dschg_day_diff,
                                             TRUE ~ 180))
df_gab_comp_age_2 = df_gab_comp_age_2 %>% mutate(readm_time = case_when(Readm_180_Init==1 ~
readm_day_diff-dschg_day_diff,
                                              Mort_180_Init==1 ~ death_day_diff-dschg_day_diff,
                                              TRUE ~ 180),
                                 mort_time = case_when(Mort_180_Init==1 ~ death_day_diff-dschg_day_diff,
                                             TRUE ~ 180))
df_gab_comp_age_3 = df_gab_comp_age_3 %>% mutate(readm_time = case_when(Readm_180_Init==1 ~
readm_day_diff-dschg_day_diff,
                                              Mort_180_Init==1 ~ death_day_diff-dschg_day_diff,
                                              TRUE ~ 180),
                                 mort_time = case_when(Mort_180_Init==1 ~ death_day_diff-dschg_day_diff,
                                             TRUE ~ 180))

##Make race and comorbidities into indicator variables

df_gab_comp_age_1 = df_gab_comp_age_1 %>%
  mutate(race_white = as.numeric(race=="White"),
      race_asian = as.numeric(race=="Asian"),
      race_black = as.numeric(race=="Black"),
      race_hispanic = as.numeric(race=="Hispanic"),
      race_native = as.numeric(race=="North American Native"),
      race_other = as.numeric(race=="Other"),
      region_ne = case_when(is.na(USRegions)~0,
                TRUE~as.numeric(USRegions=="Northeast")),
      dem_ind = as.numeric(dementia=="Yes"),
      cvd_ind = as.numeric(cvd=="Yes"),
      ulc_ind = as.numeric(ulcers=="Yes"))

df_gab_comp_age_2 = df_gab_comp_age_2 %>%
  mutate(race_white = as.numeric(race=="White"),
      race_asian = as.numeric(race=="Asian"),
      race_black = as.numeric(race=="Black"),
      race_hispanic = as.numeric(race=="Hispanic"),
      race_native = as.numeric(race=="North American Native"),
      race_other = as.numeric(race=="Other"),
      region_ne = case_when(is.na(USRegions)~0,
                TRUE~as.numeric(USRegions=="Northeast")),
      dem_ind = as.numeric(dementia=="Yes"),
      cvd_ind = as.numeric(cvd=="Yes"),
      ulc_ind = as.numeric(ulcers=="Yes"))
```

```
df_gab_comp_age_3 = df_gab_comp_age_3 %>%
  mutate(race_white = as.numeric(race=="White"),
         race_asian = as.numeric(race=="Asian"),
         race_black = as.numeric(race=="Black"),
         race_hispanic = as.numeric(race=="Hispanic"),
         race_native = as.numeric(race=="North American Native"),
         race_other = as.numeric(race=="Other"),
         region_ne = case_when(is.na(USRegions)~0,
                          TRUE~as.numeric(USRegions=="Northeast")),
         dem_ind = as.numeric(dementia=="Yes"),
         cvd_ind = as.numeric(cvd=="Yes"),
         ulc_ind = as.numeric(ulcers=="Yes"))

#Nearest Matching
m.out_comp_age_1 <- matchit(med_type ~ sex + race_white + dschg_day_diff + mrs_predictions, data =
df_gab_comp_age_1,
                   method = "nearest", distance = "mahalanobis", ratio = 5)
m.out_comp_age_2 <- matchit(med_type ~ sex + race_white + dschg_day_diff + mrs_predictions, data =
df_gab_comp_age_2,
                    method = "nearest", distance = "mahalanobis", ratio = 5)
m.out_comp_age_3 <- matchit(med_type ~ sex + race_white + dschg_day_diff + mrs_predictions, data =
df_gab_comp_age_3,
                        method = "nearest", distance = "mahalanobis", ratio = 4)
#Exact Matching
m.out_comp_age_1 <- matchit(med_type ~ dschg_day_diff, data = df_gab_comp_age_1,
                   method = "exact")
m.out_comp_age_2 <- matchit(med_type ~ dschg_day_diff, data = df_gab_comp_age_2,
                    method = "exact")
m.out_comp_age_3 <- matchit(med_type ~ dschg_day_diff, data = df_gab_comp_age_3,
                        method = "exact")

##Data frames
df_surv_comp_age_1 <- match.data(m.out_comp_age_1)
df_surv_comp_age_2 <- match.data(m.out_comp_age_2)
df_surv_comp_age_3 <- match.data(m.out_comp_age_3)

##Correcting exceptions (same time of readmission and death)

#Patients with readmission and mortality on the same day
same_vec =
which(df_surv_comp_age_1$Readm_180_Init[df_surv_comp_age_1$readm_time==df_surv_comp_age_1$mort_t
ime]==1)

pat_vec = which(df_surv_comp_age_1$readm_time==df_surv_comp_age_1$mort_time)[same_vec]

#Add offset for mortality
df_surv_comp_age_1$mort_time[pat_vec] = df_surv_comp_age_1$readm_time[pat_vec]+0.5
```

```
same_vec =
which(df_surv_comp_age_2$Readm_180_Init[df_surv_comp_age_2$readm_time==df_surv_comp_age_2$mort_t
ime]==1)

pat_vec = which(df_surv_comp_age_2$readm_time==df_surv_comp_age_2$mort_time)[same_vec]

df_surv_comp_age_2$mort_time[pat_vec] = df_surv_comp_age_2$readm_time[pat_vec]+0.5

same_vec =
which(df_surv_comp_age_3$Readm_180_Init[df_surv_comp_age_3$readm_time==df_surv_comp_age_3$mort_t
ime]==1)

pat_vec = which(df_surv_comp_age_3$readm_time==df_surv_comp_age_3$mort_time)[same_vec]

df_surv_comp_age_3$mort_time[pat_vec] = df_surv_comp_age_3$readm_time[pat_vec]+0.5

##Add indicators for sex and recode med type

df_surv_comp_age_1 = df_surv_comp_age_1 %>%
  mutate(sex_ind = as.numeric(sex=="Female"),
         med_type = 1 - med_type)

df_surv_comp_age_2 = df_surv_comp_age_2 %>%
  mutate(sex_ind = as.numeric(sex=="Female"),
         med_type = 1 - med_type)

df_surv_comp_age_3 = df_surv_comp_age_3 %>%
  mutate(sex_ind = as.numeric(sex=="Female"),
         med_type = 1 - med_type)

##Adjust invalid survival times

df_surv_comp_age_1$readm_time[df_surv_comp_age_1$readm_time==0]=0.5

df_surv_comp_age_2$readm_time[df_surv_comp_age_2$readm_time==0]=0.5

df_surv_comp_age_3$readm_time[df_surv_comp_age_3$readm_time==0]=0.5

df_surv_comp_age_1$mort_time[df_surv_comp_age_1$mort_time==0]=0.5

df_surv_comp_age_2$mort_time[df_surv_comp_age_2$mort_time==0]=0.5

df_surv_comp_age_3$mort_time[df_surv_comp_age_3$mort_time==0]=0.5


##Mortality analysis

model_asm_age_1 = coxph(Surv(mort_time, Mort_180_Init)~med_type + dschg_day_diff + mrs_predictions, data
= df_surv_comp_age_1)
```

```
summary(model_asm_age_1)

model_asm_age_2 = coxph(Surv(mort_time, Mort_180_Init)~med_type + dschg_day_diff + mrs_predictions, data
= df_surv_comp_age_2)

summary(model_asm_age_2)

model_asm_age_3 = coxph(Surv(mort_time, Mort_180_Init)~med_type + dschg_day_diff + mrs_predictions, data
= df_surv_comp_age_3)

summary(model_asm_age_3)

##Readmission analysis

model_asm_age_1 = coxph(Surv(readm_time, Readm_180_Init)~sex_ind + dem_ind + race_white + med_type +
mrs_predictions, data = df_surv_comp_age_1, weights = weights)

summary(model_asm_age_1)

model_asm_age_2 = coxph(Surv(readm_time, Readm_180_Init)~sex_ind + dem_ind + race_white + med_type +
mrs_predictions, data = df_surv_comp_age_2, weights = weights)

summary(model_asm_age_2)

model_asm_age_3 = coxph(Surv(readm_time, Readm_180_Init)~sex_ind + dem_ind + race_white + med_type +
mrs_predictions, data = df_surv_comp_age_3, weights = weights)

summary(model_asm_age_3)

##Testing with a semi-competing risks model
#Unadjusted
form <- Formula::Formula(readm_time + Readm_180_Init | mort_time + Mort_180_Init ~ med_type | med_type |
med_type)
#Adjusted
form <- Formula::Formula(readm_time + Readm_180_Init | mort_time + Mort_180_Init ~ sex_ind + dem_ind +
race_white + med_type + mrs_predictions + dschg_day_diff | sex_ind + race_white + med_type + mrs_predictions
+ dschg_day_diff | sex_ind + race_white + med_type + mrs_predictions + dschg_day_diff)

fit_comp_age_1 = FreqID_HReg2(Formula = form, data = df_surv_comp_age_1, model = "semi-Markov",
extra_starts = 0, hazard = "weibull", frailty = TRUE, optim_method = c("BFGS"))

fit_comp_age_2 = FreqID_HReg2(Formula = form, data = df_surv_comp_age_2, model = "semi-Markov",
extra_starts = 0, hazard = "weibull", frailty = TRUE, optim_method = c("BFGS"))

fit_comp_age_3 = FreqID_HReg2(Formula = form, data = df_surv_comp_age_3, model = "semi-Markov",
extra_starts = 0, hazard = "weibull", frailty = TRUE, optim_method = c("BFGS"))

summary(fit_comp_age_3)
```

```
#### Subsetting with dementia ####

##Comparison between gabapentin and other pain meds (other pain meds are cases)
df_gab_comp_dem = df_gab%>%
  filter(med_type>0, dementia=="Yes")%>%
  mutate(med_type=1-as.integer(med_type/2))

df_gab_comp_no_dem = df_gab%>%
  filter(med_type>0, dementia=="No")%>%
  mutate(med_type=1-as.integer(med_type/2))

df_gab_comp_dem = df_gab_comp_dem %>% mutate(readm_time = case_when(Readm_180_Init==1 ~
readm_day_diff-dschg_day_diff,
                                                              Mort_180_Init==1 ~ death_day_diff-dschg_day_diff,
                                                              TRUE ~ 180),
                                          mort_time = case_when(Mort_180_Init==1 ~ death_day_diff-dschg_day_diff,
                                                             TRUE ~ 180))
df_gab_comp_no_dem = df_gab_comp_no_dem %>% mutate(readm_time = case_when(Readm_180_Init==1 ~
readm_day_diff-dschg_day_diff,
                                                                    Mort_180_Init==1 ~ death_day_diff-dschg_day_diff,
                                                                    TRUE ~ 180),
                                                mort_time = case_when(Mort_180_Init==1 ~ death_day_diff-dschg_day_diff,
                                                                   TRUE ~ 180))

##Make race and comorbidities into indicator variables

df_gab_comp_dem = df_gab_comp_dem %>%
  mutate(race_white = as.numeric(race=="White"),
         race_asian = as.numeric(race=="Asian"),
         race_black = as.numeric(race=="Black"),
         race_hispanic = as.numeric(race=="Hispanic"),
         race_native = as.numeric(race=="North American Native"),
         race_other = as.numeric(race=="Other"),
         region_ne = case_when(is.na(USRegions)~0,
                      TRUE~as.numeric(USRegions=="Northeast")),
         dem_ind = as.numeric(dementia=="Yes"),
         cvd_ind = as.numeric(cvd=="Yes"),
         ulc_ind = as.numeric(ulcers=="Yes"))

df_gab_comp_no_dem = df_gab_comp_no_dem %>%
  mutate(race_white = as.numeric(race=="White"),
         race_asian = as.numeric(race=="Asian"),
         race_black = as.numeric(race=="Black"),
         race_hispanic = as.numeric(race=="Hispanic"),
         race_native = as.numeric(race=="North American Native"),
         race_other = as.numeric(race=="Other"),
         region_ne = case_when(is.na(USRegions)~0,
                      TRUE~as.numeric(USRegions=="Northeast")),
         dem_ind = as.numeric(dementia=="Yes"),
```

```
        cvd_ind = as.numeric(cvd=="Yes"),
        ulc_ind = as.numeric(ulcers=="Yes"))

#Nearest Matching
m.out_comp_dem <- matchit(med_type ~ sex + race_white + dschg_day_diff + mrs_predictions, data =
df_gab_comp_dem,
                    method = "nearest", distance = "mahalanobis", ratio = 2)
m.out_comp_no_dem <- matchit(med_type ~ sex + race_white + dschg_day_diff + mrs_predictions, data =
df_gab_comp_no_dem,
                    method = "nearest", distance = "mahalanobis", ratio = 5)
#Exact Matching
m.out_comp_dem <- matchit(med_type ~ dschg_day_diff, data = df_gab_comp_dem,
                    method = "exact")
m.out_comp_no_dem <- matchit(med_type ~ dschg_day_diff, data = df_gab_comp_no_dem,
                    method = "exact")

##Data frames
df_surv_comp_dem <- match.data(m.out_comp_dem)
df_surv_comp_no_dem <- match.data(m.out_comp_no_dem)

##Correcting exceptions (same time of readmission and death)

#Patients with readmission and mortality on the same day
same_vec =
which(df_surv_comp_dem$Readm_180_Init[df_surv_comp_dem$readm_time==df_surv_comp_dem$mort_time]
==1)

pat_vec = which(df_surv_comp_dem$readm_time==df_surv_comp_dem$mort_time)[same_vec]

#Add offset for mortality
df_surv_comp_dem$mort_time[pat_vec] = df_surv_comp_dem$readm_time[pat_vec]+0.5

same_vec =
which(df_surv_comp_no_dem$Readm_180_Init[df_surv_comp_no_dem$readm_time==df_surv_comp_no_dem$
mort_time]==1)

pat_vec = which(df_surv_comp_no_dem$readm_time==df_surv_comp_no_dem$mort_time)[same_vec]

df_surv_comp_no_dem$mort_time[pat_vec] = df_surv_comp_no_dem$readm_time[pat_vec]+0.5

##Add indicators for sex and recode med type

df_surv_comp_dem = df_surv_comp_dem %>%
  mutate(sex_ind = as.numeric(sex=="Female"),
        med_type = 1 - med_type)

df_surv_comp_no_dem = df_surv_comp_no_dem %>%
  mutate(sex_ind = as.numeric(sex=="Female"),
        med_type = 1 - med_type)
```

```
##Adjust invalid survival times

df_surv_comp_dem$readm_time[df_surv_comp_dem$readm_time==0]=0.5

df_surv_comp_no_dem$readm_time[df_surv_comp_no_dem$readm_time==0]=0.5

df_surv_comp_dem$mort_time[df_surv_comp_dem$mort_time==0]=0.5

df_surv_comp_no_dem$mort_time[df_surv_comp_no_dem$mort_time==0]=0.5

##Mortality analysis

model_asm_no_dem = coxph(Surv(mort_time, Mort_180_Init)~med_type + dschg_day_diff + mrs_predictions, data = df_surv_comp_no_dem)

summary(model_asm_no_dem)

model_asm_dem = coxph(Surv(mort_time, Mort_180_Init)~med_type + dschg_day_diff + mrs_predictions, data = df_surv_comp_dem)

summary(model_asm_dem)

##Readmission analysis

model_asm_no_dem = coxph(Surv(readm_time, Readm_180_Init)~age + sex_ind + race_white + med_type + mrs_predictions, data = df_surv_comp_no_dem, weights = weights)

summary(model_asm_no_dem)

model_asm_dem = coxph(Surv(readm_time, Readm_180_Init)~age + sex_ind + race_white + med_type + mrs_predictions, data = df_surv_comp_dem, weights = weights)

summary(model_asm_dem)

##Testing with a semi-competing risks model
#Unadjusted
form <- Formula::Formula(readm_time + Readm_180_Init | mort_time + Mort_180_Init ~ med_type | med_type | med_type)
#Adjusted
form <- Formula::Formula(readm_time + Readm_180_Init | mort_time + Mort_180_Init ~ age + sex_ind + race_white + med_type + mrs_predictions + dschg_day_diff | age + sex_ind + race_white + med_type + mrs_predictions + dschg_day_diff | age + sex_ind + race_white + med_type + mrs_predictions + dschg_day_diff)

fit_comp_dem = FreqID_HReg2(Formula = form, data = df_surv_comp_dem, model = "semi-Markov", extra_starts = 0, hazard = "weibull", frailty = TRUE, optim_method = c("BFGS"))

fit_comp_no_dem = FreqID_HReg2(Formula = form, data = df_surv_comp_no_dem, model = "semi-Markov", extra_starts = 0, hazard = "weibull", frailty = TRUE, optim_method = c("BFGS"))

summary(fit_comp_no_dem)
```

```
#### Subsetting with MRSScore ####

##Comparison between gabapentin and other pain meds (other pain meds are cases)
df_gab_comp_low_mrs = df_gab%>%
  filter(med_type>0, mrs_predictions==0)%>%
  mutate(med_type=1-as.integer(med_type/2))

df_gab_comp_high_mrs = df_gab%>%
  filter(med_type>0, mrs_predictions==1)%>%
  mutate(med_type=1-as.integer(med_type/2))

df_gab_comp_low_mrs = df_gab_comp_low_mrs %>% mutate(readm_time = case_when(Readm_180_Init==1 ~
readm_day_diff-dschg_day_diff,
                                                  Mort_180_Init==1 ~ death_day_diff-dschg_day_diff,
                                                  TRUE ~ 180),
                                   mort_time = case_when(Mort_180_Init==1 ~ death_day_diff-dschg_day_diff,
                                                 TRUE ~ 180))
df_gab_comp_high_mrs = df_gab_comp_high_mrs %>% mutate(readm_time = case_when(Readm_180_Init==1 ~
readm_day_diff-dschg_day_diff,
                                                   Mort_180_Init==1 ~ death_day_diff-dschg_day_diff,
                                                   TRUE ~ 180),
                                    mort_time = case_when(Mort_180_Init==1 ~ death_day_diff-dschg_day_diff,
                                                  TRUE ~ 180))

##Make race and comorbidities into indicator variables

df_gab_comp_low_mrs = df_gab_comp_low_mrs %>%
  mutate(race_white = as.numeric(race=="White"),
         race_asian = as.numeric(race=="Asian"),
         race_black = as.numeric(race=="Black"),
         race_hispanic = as.numeric(race=="Hispanic"),
         race_native = as.numeric(race=="North American Native"),
         race_other = as.numeric(race=="Other"),
         region_ne = case_when(is.na(USRegions)~0,
                     TRUE~as.numeric(USRegions=="Northeast")),
         dem_ind = as.numeric(dementia=="Yes"),
         cvd_ind = as.numeric(cvd=="Yes"),
         ulc_ind = as.numeric(ulcers=="Yes"))

df_gab_comp_high_mrs = df_gab_comp_high_mrs %>%
  mutate(race_white = as.numeric(race=="White"),
         race_asian = as.numeric(race=="Asian"),
         race_black = as.numeric(race=="Black"),
         race_hispanic = as.numeric(race=="Hispanic"),
         race_native = as.numeric(race=="North American Native"),
         race_other = as.numeric(race=="Other"),
         region_ne = case_when(is.na(USRegions)~0,
                     TRUE~as.numeric(USRegions=="Northeast")),
```

```
        dem_ind = as.numeric(dementia=="Yes"),
        cvd_ind = as.numeric(cvd=="Yes"),
        ulc_ind = as.numeric(ulcers=="Yes"))


#Nearest Matching
m.out_comp_low_mrs <- matchit(med_type ~ sex + race_white + dschg_day_diff + mrs_predictions, data = 
df_gab_comp_low_mrs,
                      method = "nearest", distance = "mahalanobis", ratio = 4)
m.out_comp_high_mrs <- matchit(med_type ~ sex + race_white + dschg_day_diff + mrs_predictions, data = 
df_gab_comp_high_mrs,
                       method = "nearest", distance = "mahalanobis", ratio = 5)
#Exact Matching
m.out_comp_low_mrs <- matchit(med_type ~ dschg_day_diff, data = df_gab_comp_low_mrs,
                      method = "exact")
m.out_comp_high_mrs <- matchit(med_type ~ dschg_day_diff, data = df_gab_comp_high_mrs,
                       method = "exact")

##Data frames
df_surv_comp_low_mrs <- match.data(m.out_comp_low_mrs)
df_surv_comp_high_mrs <- match.data(m.out_comp_high_mrs)

##Correcting exceptions (same time of readmission and death)

#Patients with readmission and mortality on the same day
same_vec = 
which(df_surv_comp_low_mrs$Readm_180_Init[df_surv_comp_low_mrs$readm_time==df_surv_comp_low_mrs
$mort_time]==1)

pat_vec = which(df_surv_comp_low_mrs$readm_time==df_surv_comp_low_mrs$mort_time)[same_vec]

#Add offset for mortality
df_surv_comp_low_mrs$mort_time[pat_vec] = df_surv_comp_low_mrs$readm_time[pat_vec]+0.5

same_vec = 
which(df_surv_comp_high_mrs$Readm_180_Init[df_surv_comp_high_mrs$readm_time==df_surv_comp_high_m
rs$mort_time]==1)

pat_vec = which(df_surv_comp_high_mrs$readm_time==df_surv_comp_high_mrs$mort_time)[same_vec]

df_surv_comp_high_mrs$mort_time[pat_vec] = df_surv_comp_high_mrs$readm_time[pat_vec]+0.5

##Add indicators for sex and recode med type

df_surv_comp_low_mrs = df_surv_comp_low_mrs %>%
  mutate(sex_ind = as.numeric(sex=="Female"),
        med_type = 1 - med_type)

df_surv_comp_high_mrs = df_surv_comp_high_mrs %>%
  mutate(sex_ind = as.numeric(sex=="Female"),
```

```
    med_type = 1 - med_type)

##Adjust invalid survival times

df_surv_comp_low_mrs$readm_time[df_surv_comp_low_mrs$readm_time==0]=0.5

df_surv_comp_high_mrs$readm_time[df_surv_comp_high_mrs$readm_time==0]=0.5

df_surv_comp_low_mrs$mort_time[df_surv_comp_low_mrs$mort_time==0]=0.5

df_surv_comp_high_mrs$mort_time[df_surv_comp_high_mrs$mort_time==0]=0.5

##Mortality analysis

model_asm_high_mrs = coxph(Surv(mort_time, Mort_180_Init)~med_type + dschg_day_diff + mrs_predictions,
data = df_surv_comp_high_mrs)

summary(model_asm_high_mrs)

model_asm_low_mrs = coxph(Surv(mort_time, Mort_180_Init)~med_type + dschg_day_diff + mrs_predictions,
data = df_surv_comp_low_mrs)

summary(model_asm_low_mrs)

##Readmission analysis

model_asm_high_mrs = coxph(Surv(readm_time, Readm_180_Init)~age + sex_ind + dem_ind + race_white +
med_type, data = df_surv_comp_high_mrs, weights = weights)

summary(model_asm_high_mrs)

model_asm_low_mrs = coxph(Surv(readm_time, Readm_180_Init)~age + sex_ind + dem_ind + race_white +
med_type, data = df_surv_comp_low_mrs, weights = weights)

summary(model_asm_low_mrs)

##Testing with a semi-competing risks model
#Unadjusted
form <- Formula::Formula(readm_time + Readm_180_Init | mort_time + Mort_180_Init ~ med_type | med_type |
med_type)
#Adjusted
form <- Formula::Formula(readm_time + Readm_180_Init | mort_time + Mort_180_Init ~ med_type +
dschg_day_diff | med_type + dschg_day_diff | med_type + dschg_day_diff)

fit_comp_low_mrs = FreqID_HReg2(Formula = form, data = df_surv_comp_low_mrs, model = "semi-Markov",
extra_starts = 0, hazard = "weibull", frailty = TRUE, optim_method = c("BFGS"))

fit_comp_high_mrs = FreqID_HReg2(Formula = form, data = df_surv_comp_high_mrs, model = "semi-Markov",
extra_starts = 0, hazard = "weibull", frailty = TRUE, optim_method = c("BFGS"))
```

```
summary(fit_comp_high_mrs)
```
